# Hybrid digital intervention cohort in university students: feasibility pilot study using smartphone- and smartwatch-based monitoring and ecological momentary interventions

**DOI:** 10.64898/2026.04.28.26351917

**Authors:** Mingyue Chen, Madlene Movia, Xin Hui Chua, Sarah Yi Xuan Tan, Shenglin Zheng, Kaiyi Jin, Thitikorn Topothai, Natarajan Padmapriya, Sarah Edney, Falk Müller-Riemenschneider

**Author notes:** These two authors contributed equally to this work (Sarah Edney, Falk Müller-Riemenschneider).

## Abstract

**Background:** University students often struggle to maintain healthy sleep, physical activity, and screen usage due to academic pressures and irregular schedules. Ecological momentary assessments (EMAs) and interventions (EMIs) offer real-time, context-aware opportunities to monitor and promote healthier behaviors. This pilot study aimed to evaluate the feasibility of a hybrid study design combining continuous monitoring with sequential randomized controlled trials (RCTs) evaluating EMIs targeting movement behaviors among university students.

**Methods:** The MOVE@NUS pilot study (September 2024 - January 2025) embedded three sequential RCTs, each targeting one behavior: sleep, physical activity, or screen time. For each RCT, participants were randomized on a 1:1:1 schedule (control, intervention 1, intervention 2). Eligible participants were first-year undergraduates, aged 18–25 years, who regularly used an iPhone and an Apple Watch. Smartwatches (primary) and smartphones (supplementary) passively and continuously tracked behaviors. EMAs (eight 3-day bursts) and web-based surveys captured self-reported behaviors and participant experience. All assessments were self-administered, and no provider assistance was involved.

**Results:** Of 229 students who met screening criteria, 65 enrolled (mean age 20.4 ± 1.5 years; 53.8% female). Questionnaire completion was high (baseline: 100.0%, midway: 89.2%, endpoint: 86.2%). EMA engagement decreased from 88.7% (first burst) to 49.2% (final burst). Passively monitored data were obtained from 62 participants (95.4%) with a mean tracking duration of 67.8 days (range: 11 to 114). Data completeness was highest for passively captured measures of physical activity, while more participant-dependent measures, such as manually uploading screen time screenshots, showed greater attrition. Overall satisfaction was 78.9% for sleep, 70.6% for physical activity, and 60.0% for screen time.

**Conclusions:** This hybrid study design is feasible and acceptable among university students, with successful integration of self-reports and passive tracking. Variations in engagement and data completeness highlight areas for optimization in future large-scale digital cohort studies.

**Trial registration:** ClinicalTrials.gov ID NCT06597890 First Posted: 2024-09-19.

## INTRODUCTION

The rapid advancement of smartphones and wearable devices has created new opportunities for delivering scalable, personalized preventive health interventions in real-world settings [1,2]. These ubiquitous devices enable continuous passive monitoring and delivery of tailored interventions based on real-time behavioral and contextual data [1,2]. Such digital approaches address the critical limitations of traditional health promotion methods, which rely on infrequent contact and retrospective self-reporting, thereby improving both the temporal granularity and ecological validity of measurement and intervention delivery [1,2].

Ecological momentary assessments and ecological momentary interventions represent key advances in leveraging advanced technological capabilities. EMAs capture real-time self-reports of behaviors, moods, and contextual factors, reducing recall bias and providing temporally granular datasets for understanding the behavioral dynamics [3,4]. EMIs build upon EMAs by delivering context-sensitive, personalized prompts that can be either static (based on baseline characteristics) or dynamically adapted using real-time data [4,5]. This approach may increase the relevance and timeliness of behavior change support [6,7]. With the integration of behavioral tracking features in smartphones and wearable devices, EMIs have the potential to transform digital health interventions [8,9].

University students are a critical population for digital health research due to their unique developmental stage, heightened vulnerability to health challenges, and heavy reliance on digital devices. [10]. Despite being perceived as a relatively healthy population, research suggests that cardiovascular risk factors and subclinical disease may emerge during this period [11]. Moreover, this life stage is characterized by academic stress, irregular schedules, increased independence, and higher social commitments, which together contribute to insufficient physical activity, inadequate sleep, and excessive recreational screen time [12–14]. These behaviors are interrelated and critically influence students’ physical and mental health outcomes [12,15–19]. In Singapore, where smartphone penetration among young adults is 100%, university students represent a tech-savvy and accessible population for testing real-time, multi-behavior digital interventions [20]. However, rigorous evaluations of multi-behavior, real-time EMIs in university settings remain sparse [21] and have shown mixed results [22].

Despite the promise of these approaches, feasibility challenges remain unexplored. Trials often report declining EMA response rates over time and difficulties in sustaining engagement that must be characterized and mitigated before scaling interventions [10,23–25]. In addition, passive sensor data completeness, asynchronous or fragmented data streams, and analytic complexity pose barriers to scaling [23,26–31]. Therefore, understanding feasibility, including recruitment and representativeness, retention, data completeness, intervention delivery, and user engagement, is essential to optimize technical, behavioral, and procedural elements for larger trials.

To address this gap, the MOVE@NUS pilot study leverages a novel hybrid study design combining continuous passive smartphone- and smartwatch-based monitoring (iPhone + Apple Watch) with multiple embedded three-arm randomized controlled trials (RCTs) testing short, targeted EMIs for sleep, physical activity, and screen time [32–34]. The primary aim of this pilot was to evaluate the feasibility of implementing this hybrid design among university students, with a focus on recruitment, retention, engagement, data completeness, intervention delivery processes, and participant experience. The secondary aim was to explore the preliminary effectiveness of behavioral change interventions to inform intervention fine-tuning and technical improvements for the main MOVE@NUS intervention cohort. The findings will provide critical methodological insights into the delivery of real-time, personalized digital interventions and serve as foundational evidence to inform the development of a future full-scale program tailored to university students [34,35].

## MATERIALS AND METHODS

### Study design

The MOVE@NUS pilot study followed a five-month hybrid design (September 2024 - January 2025), combining continuous digital monitoring with three sequentially embedded RCTs, separately targeting sleep (RCT-1), physical activity (RCT-2), or screen time (RCT-3). The study protocol has been published previously [36]. For each RCT, participants were randomized on a 1:1:1 schedule (control, intervention 1, intervention 2) and were preceded and followed by at least two weeks of digital monitoring (Fig 1). Movement behaviors were tracked using Apple Watches, with supplementary data from iPhones, all retrieved via Apple HealthKit. EMAs, administered every two weeks, collected real-time self-reported data. Both EMIs and EMAs were text-based, with each session taking less than two minutes to complete. Online questionnaires were administered at baseline, midway (2.5 months), and endpoint (5 months). All assessments and intervention content were delivered via each participant’s own smartphone and smartwatch. This study adhered to the Consolidated Standards of Reporting Trials (CONSORT) 2025 updated guidelines and CONSORT-EHEALTH checklist (V.1.6.1) [37,38].

**Fig 1.**
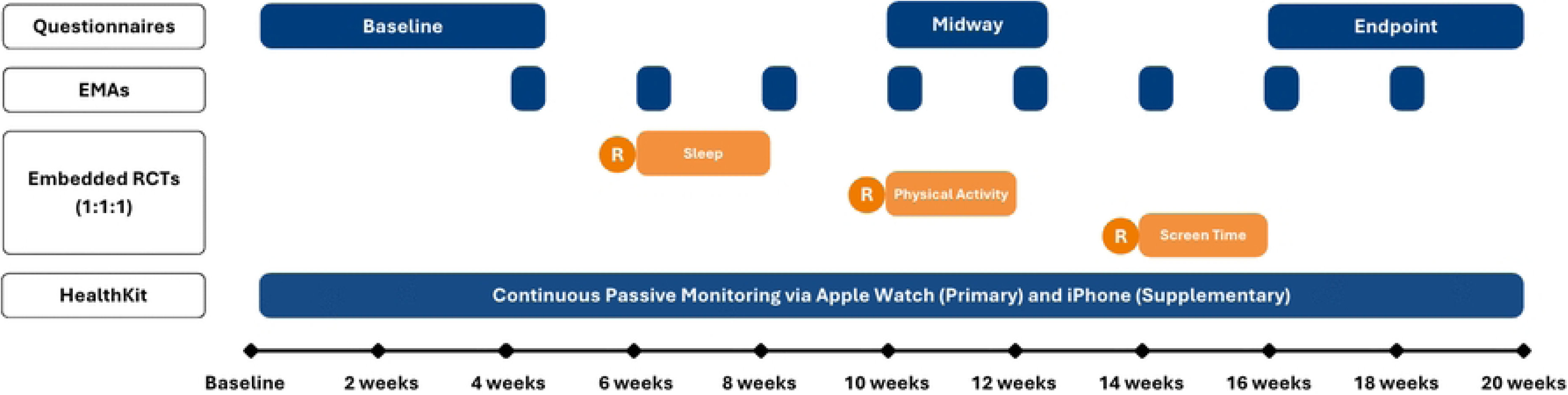
Study design of MOVE@NUS pilot study: assessments, data collection and interventions. (A)The study began with online assessments, which were also administered at midway (2.5 months), and endpoint (5 months). Ecological Momentary Assessments (EMAs) were administered in bursts throughout the study, with each burst lasting 3 days. The intervention period comprised three sequential, embedded RCTs (each lasting 2 weeks): RCT-1 (Sleep), RCT-2 (Physical Activity), and RCT-3 (Screen Time), with continuous passive monitoring via Apple Watch and iPhone. Participants were re-randomized (’R’) 1:1:1 to intervention or control groups at the start of each new RCT phase.

### IRB approval and ethical considerations

Ethics approval for this study has been obtained from the National University of Singapore Institutional Review Board (NUS-IRB) (Approval Number: NUS-IRB-2023-800).

### Participants

Eligible participants were first-year undergraduate students at the National University of Singapore, aged 18–25 years, who owned or regularly used an iPhone with constant access to the internet and an Apple Watch (Series 6/SE or newer). First-year students were chosen to pilot the study as future iterations of the study intend to track cohorts of students longitudinally (from first year through to graduation), enabling assessment of long-term effectiveness. Participants also had to be willing to download the study app and to wear their watch continuously, including during sleep. Exclusion criteria included pregnancy or planned overseas travel exceeding six weeks during the study period.

### Recruitment

Recruitment occurred through multiple channels from September 9 and October 14, 2024. The University Health Centre distributed recruitment emails to the university email addresses of all eligible students. Supplementary recruitment efforts included promotion through social media platforms commonly used by Singaporean students (Telegram, WhatsApp, and WeChat) with specific targeting of NUS freshman groups, as well as strategic deliveries of informational posters across campus. Interested students were directed to complete a brief online questionnaire to confirm their eligibility. Eligible participants were invited to attend an on-site enrollment session, during which written informed consent was obtained. For participants under the age of 21 years, consent from a legal guardian was also required. Access to HealthKit data was requested through the study-specific smartphone app, which required participants to provide consent for their data to be accessed. Participants were reimbursed at the midway and at the end of the study (S$90 in total) for their time and participation.

### Data collection

The MOVE@NUS study app was developed and tested by our research team specifically for this study using the Cogniss platform [39]. The app was integrated with HealthKit to collect movement behavior data. Unlike conventional smartphone apps that typically focus on either research prototypes or direct consumer markets, the app was designed specifically to support personalized health monitoring and behavior change interventions, capable of achieving a high level of personalization [39]. Data were collected from three main sources (Fig 1): (1) Apple Watch/iPhone HealthKit: objective and passively monitored data on sleep and PA, with minimal participant burden. Screen time data is not available via HealthKit and was therefore collected weekly as iOS Settings screenshots; (2) EMAs: Captured self-reported measures of screen use, delivered in eight bursts, each comprising three consecutive days of assessment, and repeated every two weeks. Each burst included one prompt per day, balancing data quality with participant burden. Completion of at least one daily EMA survey within a burst was considered a completed burst for analysis purposes; (3) Questionnaires: Self-reported questionnaires were administered via a REDCap (Research Electronic Data Capture) survey on participants’ smartphones at baseline, midway, and the study endpoint [40,41]. The questionnaires measure demographics, lifestyles, and movement behaviors. Completion of the full questionnaire took approximately 30 minutes. Participants who did not complete the questionnaires received a reminder email after one week, and if necessary, a second reminder was sent one week later.

### Intervention

#### MOVE@NUS: digital health behavior app

The MOVE@NUS app underwent internal quality checks before study launch to ensure system stability, usability, and reliable intervention delivery. Preliminary testing by the research team and a small pilot group helped identify interface, navigation, and scheduling issues. Based on identified issues, iterative refinements were made to visual design, page layout, assessment scheduling, and the wording of in-app messages and notifications to optimize participant experience. Intervention delivery fidelity was verified through checks on notification timing (arrival within ±10 minutes of the programmed schedule) and content accuracy (correct display of messages according to predefined triggers). These procedures ensured that EMIs were delivered consistently during the study.

The MOVE@NUS app was made exclusively available to enrolled study participants through a secure access protocol. During on-site enrollment sessions, participants accessed the platform by scanning a QR code that directed them to the registration page. Participants registered using pre-assigned usernames and passwords, after which an automated consent page for HealthKit data synchronization would be generated.

Upon providing informed consent for data access, participants gained full access to study features and monitoring capabilities.

#### Ecological momentary interventions in the MOVE@NUS pilot study

Multiple EMI formats are used, including evidence-based standard health messages, and personalized reminders derived from HealthKit data, self-reported behaviors, and stated preferences (Table 1). Interventions were selected a priori based on existing scientific evidence to serve as test cases for evaluating the technical and behavioral feasibility of delivering real-time, personalized interventions, rather than definitively establishing intervention effectiveness. To minimize notification fatigue and maintain participant engagement, the notification system was programmed with built-in limits, delivering no more than seven notifications per day during any intervention phase, regardless of group assignment.

**Table 1.** Summaries of ecological momentary interventions implemented in MOVE@NUS pilot study.

| RCT | Group | Content Description | Message Frequency |
| --- | --- | --- | --- |
| Sleep | Sleep Duration Nudge | Feedback on the past 7-day sleep duration with guideline-based recommendations | Weekly reports + Daily nudge (late evening) |
|  | Sleep Hygiene Choice | Evidence-based sleep hygiene strategies with participant selection | Weekly strategy selection + Daily nudge (late morning) |
|  | Control Group | No intervention | - |
| Physical Activity | Stair Climbing Nudge | Encouragement and contextual nudges based on real-time flights climbed to take stairs instead of elevators or escalators | Daily report (late morning) + Daily nudges (early afternoon, late afternoon) |
|  | VPA (Vigorous Physical Activity) Choice | Brief, intense physical activity options with daily selection and nudges | Daily strategy selection (late morning) + Daily nudges (early afternoon, late afternoon) |
|  | Control Group | No intervention | - |
| Screen Time | Screen Time Break Nudge | Break reminders with alternative activity suggestions | Daily report (late morning) + Daily nudges (6 total: noon to night, every ~2 hours) |
|  | Screen Time Goal-Setting | Personalized daily screen time reduction goal setting and monitoring | Daily report (late morning) + Daily nudge (afternoon) |
|  | Control Group | No intervention | - |

Following group allocation and one day before the commencement of each intervention phase, participants received an instructional WhatsApp message about the upcoming intervention. They were informed of their specific intervention assignment to maintain allocation concealment. Throughout the study duration, a dedicated project email and WhatsApp account remained available for participant inquiries and to send periodic reminders about app engagement to ensure successful data synchronization and intervention delivery.

#### Detailed intervention protocols by RCT

RCT-1: Sleep

- Intervention Group 1 - Sleep Duration Nudge: Feedback was personalized using objective sleep data and aligned with established sleep duration guidelines (7-9 hours/day) for young adults, providing individualized recommendations [42,43]. Participants received weekly feedback reports based on their previous 7-day sleep duration patterns from HealthKit data, accompanied by daily reminder messages [43].
- Intervention Group 2 - Sleep Hygiene Choice: Sleep hygiene interventions improve sleep quality in university students [44]. Through the app, participants selected weekly from a list of evidence-based sleep hygiene strategies (e.g., reducing screen use before bedtime, no coffee after 3 PM) [44]. Daily reminders were tailored to strategies actively selected by each participant, ensuring relevance and personal commitment to the chosen approaches.
- Control Group: Participants in this group continued standard app usage and data collection without receiving any intervention messages.

RCT-2: Physical Activity

- Intervention Group 1 - Stair Climbing Nudge: Stair climbing is a practical and effective physical activity leveraged existing environmental opportunities for physical activity enhancement [45,46]. Participants received daily encouragement and contextual reminders to choose stairs over elevators or escalators, with feedback based on HealthKit data on actual flights of stairs climbed.
- Intervention Group 2 - VPA (Vigorous Physical Activity) Choice: Short bursts of activity improve cardiovascular health [46,47]. Participants received daily prompts to select from a comprehensive menu of brief, vigorous-intensity physical activities (e.g., brisk walking sessions between classes, squats while waiting), designed to be integrated into typical university student routines and environments. [46,47].
- Control Group: Participants in this group continued standard app usage and data collection without receiving any intervention messages.

RCT-3: Screen Time

- Intervention Group 1 – Screen Time Break Nudge: Breaking prolonged screen use can reduce digital fatigue and promote healthier habits [48]. Participants received contextually-aware reminders to take breaks from screen use, with suggestions for brief alternative activities (e.g., stretching, brief walks) [48]. The system queried the current smartphone usage status before providing tailored break suggestions, ensuring intervention relevance.
- Intervention Group 2 - Screen Time Goal-Setting: Personalized goal-setting interventions improve adherence to behavior change [49]. Participants set personalized daily screen time reduction goals through the app interface [49]. The intervention utilized iOS screen time data to provide daily personalized goal-setting support and progress feedback.
- Control Group: Participants in this group continued standard app usage and data collection without receiving any intervention messages.

### Outcomes

#### Feasibility

The feasibility of this study was assessed through: (1) Recruitment Metrics: eligibility rates, enrollment rates, and non-response rates. (2) Response Rates and Retention: questionnaire completion rates, EMA response rates, and HealthKit data completeness (number of days tracked, retention within each intervention phase, and continuity of tracking days). (3) Participant Experience with MOVE@NUS: Feedback was gathered through prompts at baseline and after each RCT phase, which included both Likert scales and open-text questions. To structure participant experience, this study adopts a micro–macro framework which integrates elements from established models, including the Technology Acceptance Model (TAM), COM-B (Capability, Opportunity, Motivation, Behavior), MARS (Motivation, Ability, Role Perception, Situational Factors), and social/digital determinants of health [50–53]. This framework supports systematic data collection across two levels:

- Micro level: immediate, direct experiences including intervention-specific feedback, user experience with the application, and overall study experience.
- Macro level: broader contextual factors influencing participants’ engagement.

#### Preliminary effectiveness

Preliminary effectiveness of EMIs implemented was assessed using a combination of objective and self-reported measures, selected for their relevance to each targeted behavior and their feasibility for continuous data collection in the study setting. Analyses included only participants with available data from both pre-intervention and intervention phases.

(1) Sleep (RCT-1): Sleep duration was derived from HealthKit, which records time in bed as a proxy for sleep duration. This measure was chosen because sleep duration has an important influence on many biological processes, such as inflammation, glucose regulation, appetite, and energy expenditure, as well as psychological processes such as memory consolidation and attention [54].
(2) Physical activity (RCT-2): Step counts and flights climbed were derived from HealthKit. Step counts are a widely accepted and interpretable metric of overall physical activity [55], while stair climbing captures short bouts of vigorous physical activity, which have been shown to improve cardiometabolic health in everyday settings [45]. In Singapore, it has been shown that smartphone- and wearable-based interventions demonstrated feasibility for promoting both step counts and stair climbing, suggesting their potential as physical activity targets within digital health strategies [56]. These outcomes were chosen to capture both volume and intensity of physical activity within the naturalistic campus environment.
(3) Screen time (RCT-3): Total smartphone screen time was collected from weekly uploads of iOS Settings screenshots through EMA bursts. Prolonged screen time not only displaces time for physical activity [57,58] but also disrupts sleep through blue-light exposure and heightened psychological engagement [59,60], potentially altering cognitive processing [61] and increasing overweight/obesity rates [62].

### Sample size

Given the exploratory nature of this pilot study, formal sample size calculations were not conducted. The study aimed to assess recruitment feasibility within the constraints of strict eligibility requirements and a complex embedded RCT design, with sample size determined by recruitment capacity during the study period.

### Randomization

The randomization sequence was generated using a secure, computer-based random number generator with no manual input, to avoid bias. For each RCT, participants were randomized on a 1:1:1 schedule (control, intervention 1, intervention 2). Once a participant was confirmed as eligible and provided informed consent, their treatment assignment was determined automatically based on the pre-generated randomization sequence for each respective RCT phase. The randomization occurred seamlessly within the app infrastructure, maintaining allocation concealment throughout the study period. This approach helped distribute potential carryover effects across conditions and minimized systematic bias from previous intervention assignments. While this approach could not eliminate carryover effects between sequential RCTs, it aimed to balance their influence across the study population.

### Blinding

The study employed an Assessor-Blind design with Partial Participant Masking (Masked to Comparative Arms). Due to the nature of the behavioral intervention (daily personalized nudges in intervention groups vs. no intervention in control groups), full participant blinding was not feasible, as participants might inevitably be aware of the intervention they are receiving. However, to minimize bias, the following measures were implemented: (1) Assessor Blinding: Outcome assessors and data analysts remained blinded to group allocation until the final analysis was complete. (2) Partial Participant Masking: Participants were masked to the specific study hypotheses and the exact nature of the opposing arm. They were informed that the study compares different variations of health monitoring, without explicitly knowing which arm they belong to at each RCT phase. All app messages and interactions for the intervention groups were standardized using pre-defined templates to ensure consistency across participants.

### Data analysis

Descriptive statistics were used to assess feasibility outcomes. Categorical variables were reported as frequencies and percentages, while continuous variables were summarized using means and standard deviations (SDs). Feasibility outcomes and trends over time were visualized using line plots and bar charts to highlight patterns and potential signals of change. Given the nature of real-time digital nudging design, analyses focused on within-arm changes during the intervention period, rather than post-intervention, to reflect the ecological and real-time delivery of digital nudges. Outcome mean differences between pre- and during-intervention phases are shown with corresponding 95% confidence intervals (CIs).

All statistical analyses were conducted using R version 4.2.2, with two-sided tests at a 5% significance level.

### Adverse events

Given that all assessments and interventions were delivered via participants’ own smartphones and smartwatches, potential digital interruptions or increases in device usage were considered possible minimal risks. Participants received no more than seven push notifications per day, each lasting less than two minutes, resulting in an estimated total screen time of under 15 minutes per day attributable to the study. No other adverse events or unintended effects from the trial interventions or procedures were found. Any participant-reported issues were documented and addressed promptly. Privacy breaches were not applicable, as only HealthKit data were synchronized through the study-specific app with consent, and all other personal data on their smartphones and smartwatches remained blinded or were not retrieved.

## RESULTS

### Participant Characteristics

Among the 65 participants, 35 were female (53.8%), and their mean age was 20.4 ± 1.5 years (Table 2). Most participants (66.2%) were from the Science and Engineering faculties, 18.5% from Humanities and Business, and 15.4% from Health Sciences. Nearly half (46.7%) had a monthly household income exceeding Singapore Dollar (SGD) 10,000.

**Table 2.** Baseline demographic and clinical characteristics of study participants by RCT phase and treatment arm.

| Characteristics | All participants (n=65) | RCT-1 Sleep (n=53) |  |  | RCT-2 Physical Activity (n=65) |  |  | RCT-3 Screen Time (n=65) |  |  |
| --- | --- | --- | --- | --- | --- | --- | --- | --- | --- | --- |
| RCT group (n) |  | Sleep Duration (18) | Sleep Hygiene (18) | Control (17) | Stair Climbing (22) | VPA (22) | Control (21) | Screen Time Break (22) | Screen Time Goal-Setting (22) | Control (21) |
| Age (years) | 20.4±1.5 | 20.9±1.7 | 20.7±1.8 | 19.7±1.1 | 20.2±1.5 | 20.6±1.8 | 20.3±1.2 | 20.4±1.4 | 20.7±1.5 | 20.1±1.6 |
| Sex |  |  |  |  |  |  |  |  |  |  |
| Female | 35(53.8) | 14(77.8) | 8(44.4) | 5(29.4) | 14(63.6) | 7(31.8) | 9(42.9) | 13(59.1) | 9(40.9) | 8(38.1) |
| Male | 30(46.2) | 4(22.2) | 10(55.6) | 12(70.6) | 8(36.4) | 15(68.2) | 12(57.1) | 9(40.9) | 13(59.1) | 13(61.9) |
| Ethnicity |  |  |  |  |  |  |  |  |  |  |
| Chinese | 52(80.0) | 13(72.2) | 15(83.3) | 15(88.2) | 17(77.3) | 21(95.5) | 14(66.7) | 16(72.7) | 20(90.9) | 16(76.2) |
| Others | 13(20.0) | 5(27.8) | 3(16.7) | 2(11.8) | 5(22.7) | 1(4.5) | 7(33.3) | 6(27.3) | 2(9.1) | 5(23.8) |
| Faculty |  |  |  |  |  |  |  |  |  |  |
| Humanities and Business | 12(18.5) | 2(11.1) | 4(22.2) | 6(35.3) | 1(4.6) | 4(18.2) | 7(33.3) | 4(18.2) | 4(18.2) | 4(19.1) |
| Science and Engineering | 43(66.2) | 14(77.8) | 14(77.8) | 9(52.9) | 19(86.4) | 13(59.1) | 11(52.4) | 15(68.2) | 14(63.6) | 14(66.7) |
| Health Sciences | 10(15.4) | 2(11.1) | 0(0.0) | 2(11.8) | 2(9.1) | 5(22.7) | 3(14.3) | 3(13.6) | 4(18.2) | 3(14.3) |
| Monthly household income (SGD) |  |  |  |  |  |  |  |  |  |  |
| Less than 3999 | 10(22.2) | 4(30.8) | 2(15.4) | 3(16.7) | 3(18.8) | 4(26.7) | 3(21.4) | 2(13.3) | 5(33.3) | 3(20.0) |
| 4000 to 9999 | 14(31.1) | 1(7.7) | 3(23.1) | 1(8.3) | 3(18.8) | 0(0.0) | 2(14.2) | 3(20.0) | 0(0.0) | 2(13.3) |
| More than 10000 | 21(46.7) | 8(61.5) | 8(61.5) | 8(66.7) | 10(62.5) | 11(73.3) | 9(64.3) | 10(66.7) | 10(66.7) | 10(66.7) |
| Refuse to answer | 20 | 5 | 5 | 5 | 6 | 7 | 7 | 7 | 7 | 6 |

(A) Data are presented as N (%) for categorical variables and Mean ± SD for continuous variables unless otherwise stated.
(B) Of the enrolled participants, 53 joined before RCT-1 (Enrollment Batch 1), while 12 were enrolled after RCT-1 but before RCT-2 (Enrollment Batch 2), and thus only participated in RCT-2 and RCT-3.

### Feasibility

#### Participant Recruitment

Following the email invitation, 229 students met the initial eligibility criteria based on the screening questionnaire (Fig 2), and 110 expressed interest in enrolling onsite. During onsite enrollment, 38 individuals did not attend, and 7 were excluded after onsite eligibility checks were conducted. A total of 65 participants provided informed consent and were successfully enrolled (overall enrollment rate: 28.4%). Among these, 53 were enrolled before the start of RCT-1 (Enrollment Batch 1), while 12 were enrolled before the second intervention (RCT-2) and were therefore included only in RCT-2 and RCT-3 (Enrollment Batch 2). Due to the nature of the online screening process, where eligibility criteria were embedded in the recruitment emails, and not all respondents proceeded to the full questionnaire, an accurate eligibility rate could not be calculated.

**Fig 2.**
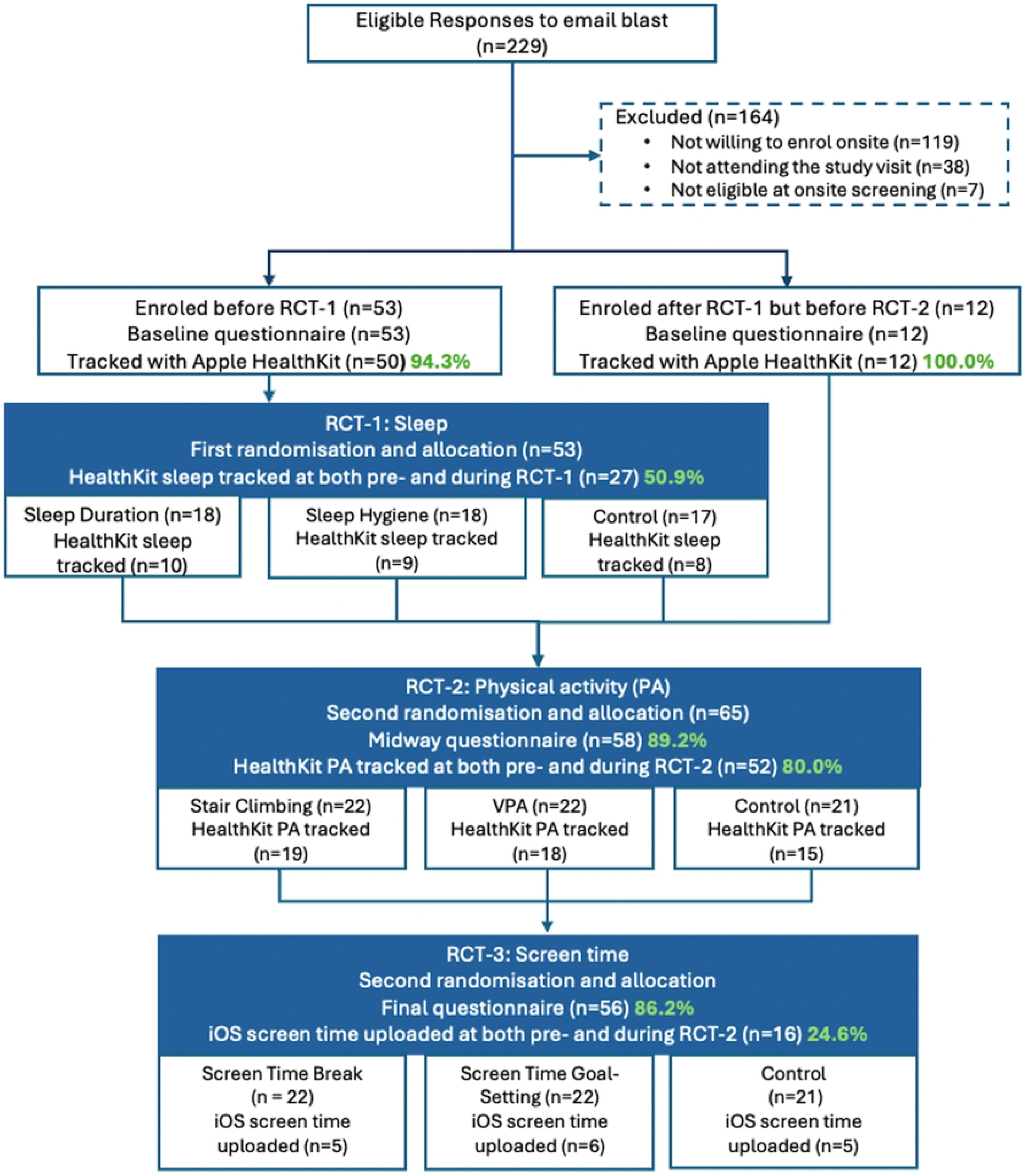
Participant flow diagram.

#### Response rates and retention

##### (1) Questionnaire response rates

65 were enrolled and completed the online baseline survey, 58 (89.2%) completed the midway online survey, and 56 (86.2%) completed the endpoint survey.

##### (2) EMA response rates

Among Enrollment Batch 1 (n=53), 47 (88.7%) completed the initial EMA burst, while Enrollment Batch 2 (n=12) completed their first EMA burst upon entry. By study conclusion, 24 of 53 (45.3%) participants and 8 of 12 (66.7%) participants completed the final EMA burst (Fig 3). Individual participant engagement varied considerably across the study period. Twenty participants (30.8%) completed all 7 EMA bursts. Additionally, 4 (6.2%) participants completed 6 bursts, 13 (20.0%) completed 5 bursts, 7 (10.8%) completed 4 bursts, 6 (9.2%) completed 3 bursts, 5 (7.7%) completed 2 bursts, and 6 (9.2%) participants completed only 1 burst.

**Fig 3.**
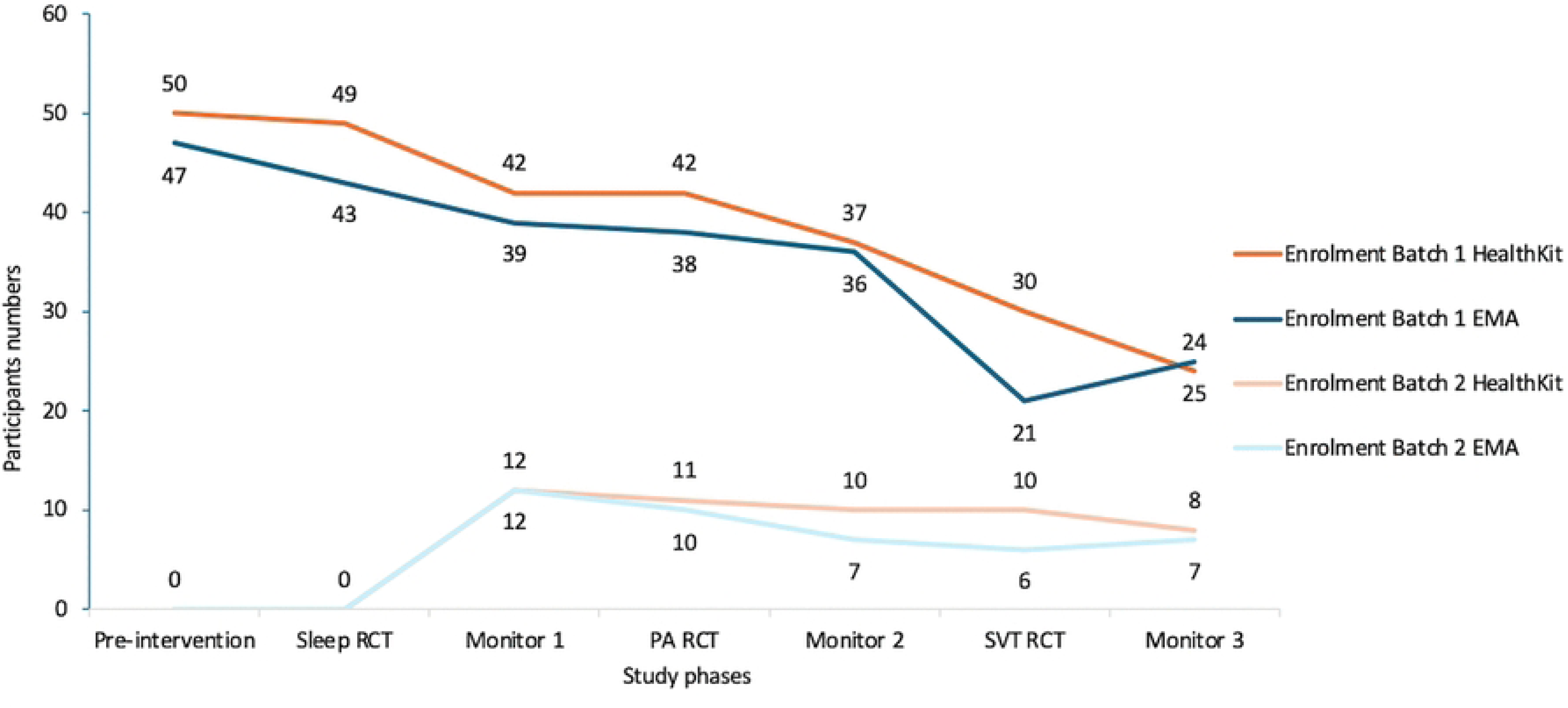
Data completeness and retention: HealthKit and EMA response rates across RCT phases (n=65). (A)Dark orange and dark blue showed participants who enrolled before RCT-1 (n=53, Enrollment Batch 1), light orange and light blue showed participants who enrolled before RCT-2 (n=12, Enrollment Batch 2)

##### (3) Apple HealthKit data completeness

The total monitoring period spanned 130 days (September 9, 2024, to January 17, 2025). HealthKit data were successfully collected from 62 of 65 (95.4%) enrolled participants, with 3 participants excluded due to delays in providing consent to access their HealthKit data during the enrollment process. The average individual tracking duration was 67.8 ± 29.1 days (range: 11 to 114 days), representing approximately 52.2% of the total possible monitoring period.

Among Enrollment Batch 1 (n=53), 50 (94.3%) provided Apple HealthKit data from enrollment, but only 24 (48.0%) continued providing data through the final monitoring phase (Fig 3). All 12 (100.0%) participants in Enrollment Batch 2 provided HealthKit data following their enrollment, and 8 (66.7%) maintained data provision through study completion. Overall, 32 of 62 participants (51.6%) who initially provided HealthKit data continued tracking to the final phase.

Fig 4 illustrates the distribution of the percentage of total days with valid HealthKit data. A substantial proportion of continuous monitoring was provided by a subset of highly engaged participants: 30 of 62 participants (48.4%) contributed 60% of the total tracked days across all participants. Data continuity varied considerably, with only 8 participants (12.9%) providing completely continuous daily data throughout their tracking period. The remaining participants experienced data gaps ranging from 1 to 32 days, primarily due to periods of app inactivity that interrupted HealthKit synchronization.

**Fig 4.**
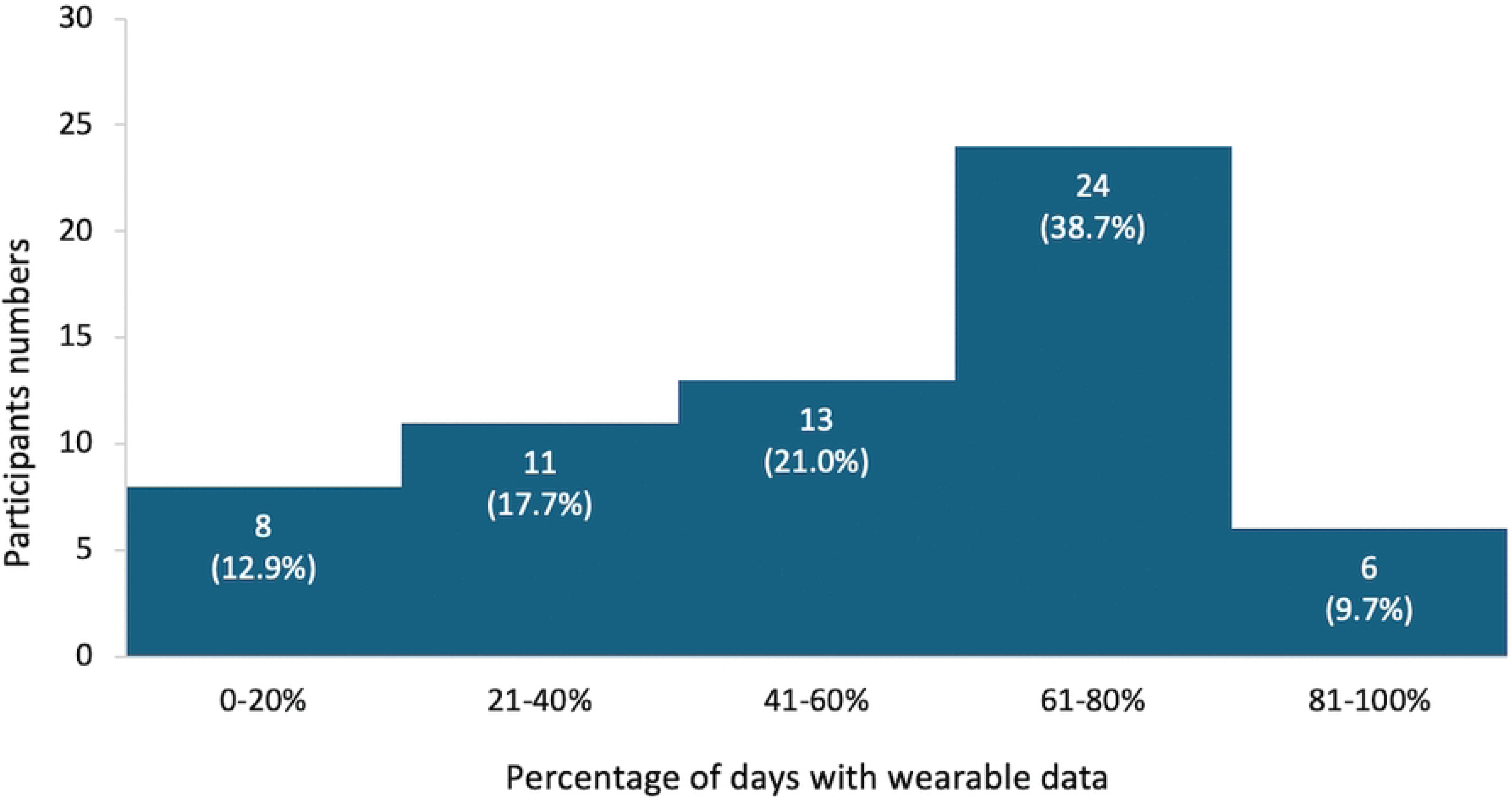
Distribution of percentage of total days with valid HealthKit data (n=62). (A)This figure presents the number of participants (y-axis) by their total percentage of total days (130 days) with valid HealthKit tracking data (x-axis), indicating data completeness. (B)HealthKit data were obtained from 62 of 65 participants; 3 were excluded due to delayed consent.

#### Participant experience

Feedback was gathered through prompts at baseline and after each RCT phase, which included both Likert scales and open-text questions. Before the RCT phases, participants generally reported high usability and comfort with the study setup. Most (85.0%) found the MOVE@NUS app easy to download and install, and to link it with their smartwatch (90.0%). Participants rated the clarity of initial study instructions (90.0%) and overall experience with setup (92.5%) positively. Comfort with using a smartwatch and smartphone apps for data collection was also high (both 80.0%), as were motivation to participate (85.0%) and confidence in the skills required to engage with the study (90.0%). Half of the participants (50.0%) reported currently using another health app.

##### (1) Micro-level feedback: usefulness, ease, and relevance

Participants generally valued the delivery of brief, low-burden behavioral nudges via mobile phones. Usefulness of strategies was reported as high for sleep (78.9%), PA (70.6%), and screen time (80.0%) (Fig 5). Participants reported that following the suggestions was easy in the PA phase (88.2%), compared to sleep (68.4%) and screen time (60.0%). Relevance to daily life was stronger for sleep (84.2%) and PA (82.4%) than for screen time (55.0%). Overall satisfaction mirrored these patterns: sleep (78.9%) and PA (70.6%), and lower for screen time (60.0%).

**Fig 5.**
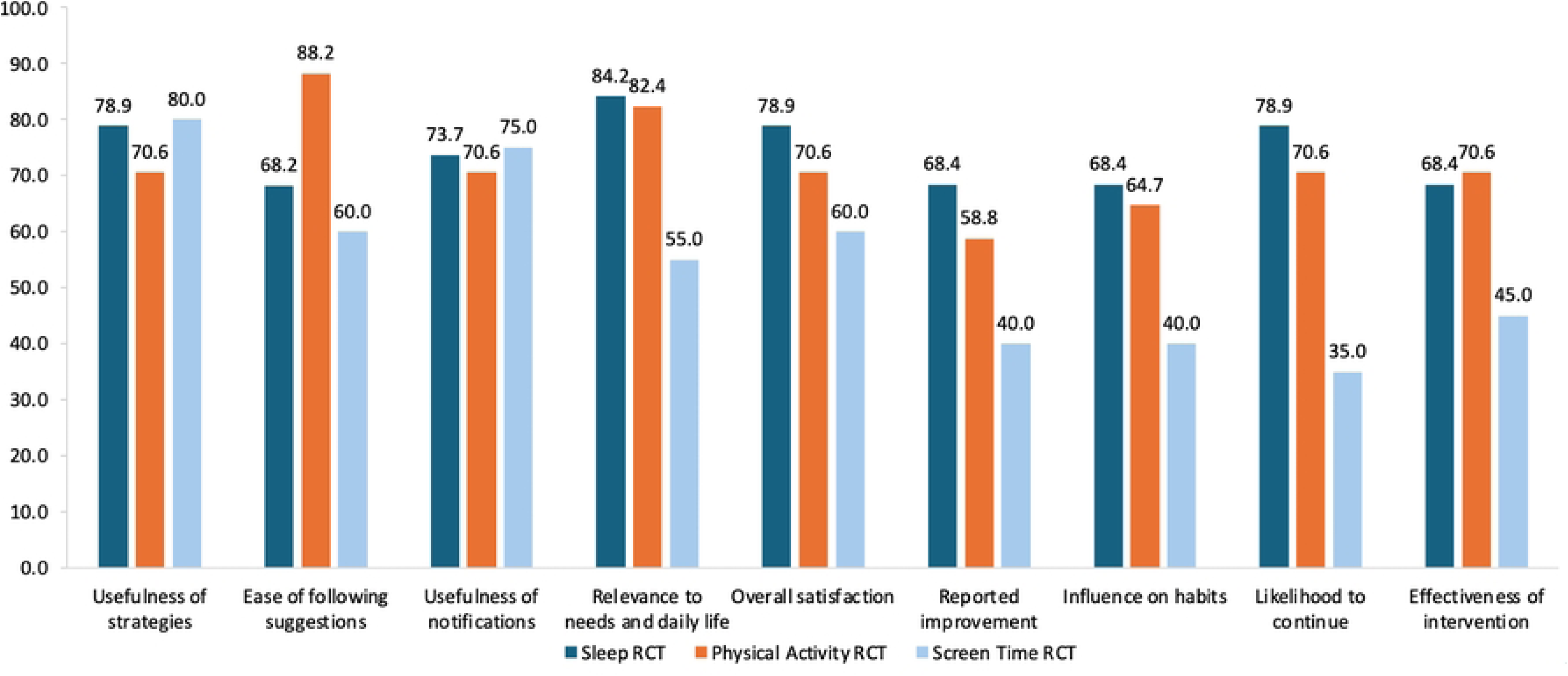
Participant experience with MOVE@NUS. (A)Data are presented as percentages responding “Yes” to all the binary questions. For the Sleep RCT (blue), 36 participants were in intervention groups, with 19 responding; for the Physical Activity RCT (orange), 44 were in intervention groups, with 17 responding; and for the Screen Time RCT (green), 44 were in intervention groups, with 20 responding; (B)The first five bar groups (Usefulness of strategies to Overall satisfaction) were micro-level outcomes, and the last four bar groups (Reported improvement to Effectiveness of intervention) were macro-level outcomes.

##### (2) Macro-level feedback: perceived impact

Around two-thirds of participants allocated to the intervention groups for the sleep interventions (68.4%) and PA interventions (58.8%) reported influences on their habits. In comparison, fewer did so in the screen time intervention groups (40.0%). Likelihood to continue using strategies was highest for sleep (78.9%), followed by PA (70.6%), and lowest for screen time (35.0%). Perceived effectiveness was moderate for sleep (68.4%) and PA (70.6%), and lower for screen time (45.0%).

##### (3) Open-text feedback

Participants highlighted that notifications acted as consistent reminders and nudges for healthy behaviors. Positive feedback emphasized simplicity, convenience, and awareness-raising.

- RCT-1: Sleep Participants noted increased awareness of their sleep patterns (*“When I key in my sleeping time, it makes me realize how bad my schedule is”*), with notifications serving as useful reminders. Challenges included academic workload barriers (*“The only sleep intervention possible is reducing my university workload”*) and notification fatigue.
- RCT-2: Physical Activity Participants described subtle behavior changes (*“Over time it crept into my subconscious that I needed to take the stairs more often”*) and appreciated the educational links provided. However, some perceived the interventions as repetitive and experienced occasional app crashes. Some participants also expressed interest in having a progress-tracking feature (*“I think the app could provide a function to track our progress. It would be more motivating as we can see our progress”*).
- RCT-3: Screen Time
- Participants found the notifications helpful in reminding them of screen time limits and offering practical strategies (*“It was useful to have the suggestions provided to me as I would not search for them by myself”*). Yet, reminders were sometimes counterproductive, as checking notifications led to additional phone use (*“The push notifications to check if I am using my phone sometimes resulted in me picking up my phone to check and respond to the notification, then continuing to use other apps, increasing my screen time”*). Participants also suggested incorporating more real-time incentives and visually engaging reminders to enhance effectiveness.

#### Preliminary Effectiveness of Interventions

Data completeness was highest for physical activity metrics (flights climbed, step counts), with 15-19 participants per group (out of 21-22 originally assigned per group) providing analyzable Apple HealthKit data (Table 3). Sleep data were available for 8-10 participants in each group (out of 21-22 originally assigned per group). In contrast, iOS screen time data (21-22 originally assigned per group), while technically passively collected, required participants to manually upload screenshots for researcher interpretation, resulting in small sample sizes (5-6 per group).

**Table 3.**
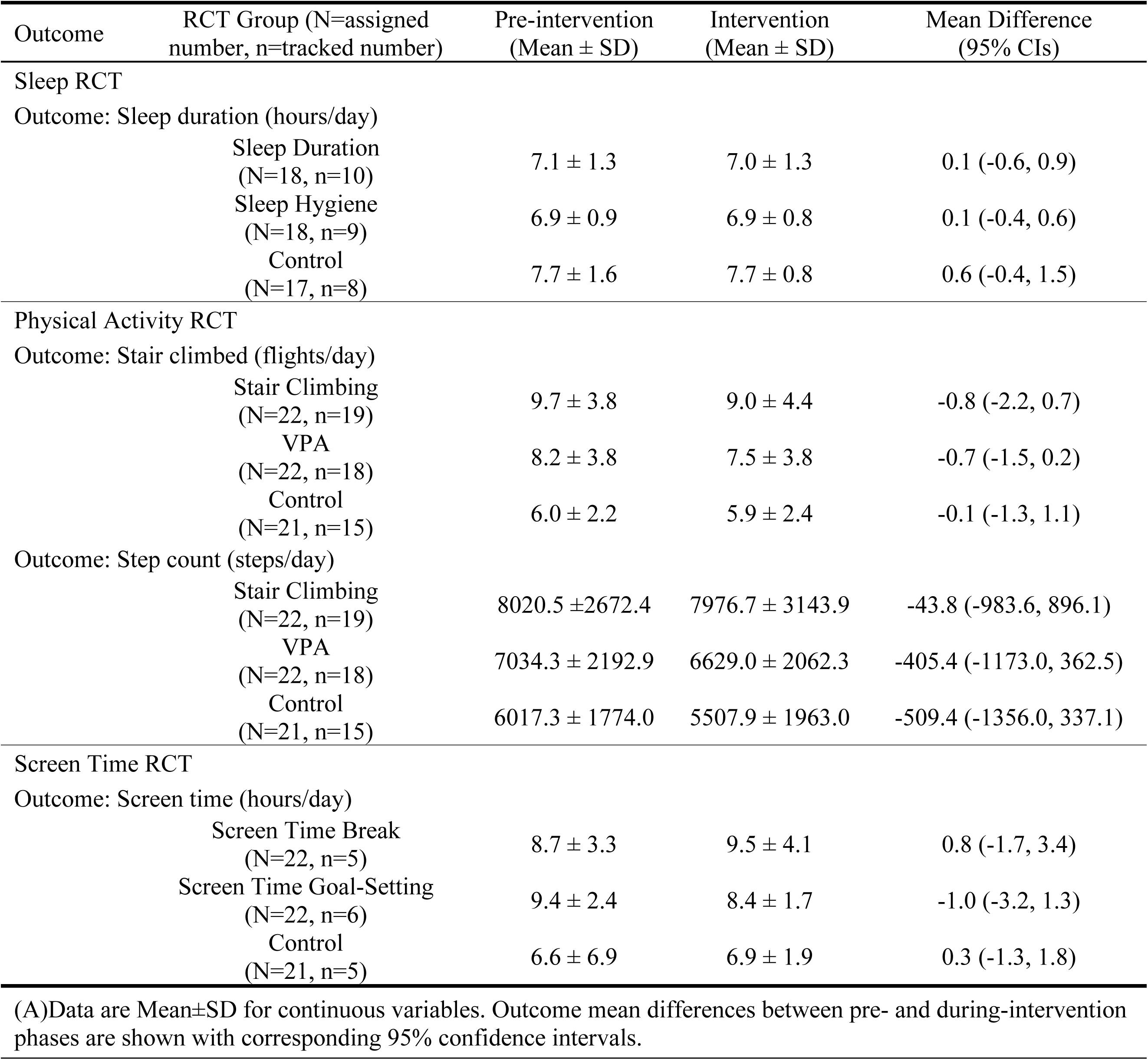
Objective health behavior outcomes from Apple HealthKit data: comparison between pre-intervention and intervention phases.

| Outcome | RCT Group (N=assigned number, n=tracked number) | Pre-intervention (Mean ± SD) | Intervention (Mean ± SD) | Mean Difference (95% CIs) |
| --- | --- | --- | --- | --- |
| Sleep RCT |  |  |  |  |
| Outcome: Sleep duration (hours/day) |  |  |  |  |
| | Sleep Duration (N=18, n=10) | 7.1 ± 1.3 | 7.0 ± 1.3 | 0.1 (-0.6, 0.9) |
| | Sleep Hygiene (N=18, n=9) | 6.9 ± 0.9 | 6.9 ± 0.8 | 0.1 (-0.4, 0.6) |
| | Control (N=17, n=8) | 7.7 ± 1.6 | 7.7 ± 0.8 | 0.6 (-0.4, 1.5) |
| Physical Activity RCT |  |  |  |  |
| Outcome: Stair climbed (flights/day) |  |  |  |  |
| | Stair Climbing (N=22, n=19) | 9.7 ± 3.8 | 9.0 ± 4.4 | -0.8 (-2.2, 0.7) |
| | VPA (N=22, n=18) | 8.2 ± 3.8 | 7.5 ± 3.8 | -0.7 (-1.5, 0.2) |
| | Control (N=21, n=15) | 6.0 ± 2.2 | 5.9 ± 2.4 | -0.1 (-1.3, 1.1) |
| Outcome: Step count (steps/day) |  |  |  |  |
| | Stair Climbing (N=22, n=19) | 8020.5 ± 2672.4 | 7976.7 ± 3143.9 | -43.8 (-983.6, 896.1) |
| | VPA (N=22, n=18) | 7034.3 ± 2192.9 | 6629.0 ± 2062.3 | -405.4 (-1173.0, 362.5) |
| | Control (N=21, n=15) | 6017.3 ± 1774.0 | 5507.9 ± 1963.0 | -509.4 (-1356.0, 337.1) |
| Screen Time RCT |  |  |  |  |
| Outcome: Screen time (hours/day) |  |  |  |  |
| | Screen Time Break (N=22, n=5) | 8.7 ± 3.3 | 9.5 ± 4.1 | 0.8 (-1.7, 3.4) |
| | Screen Time Goal-Setting (N=22, n=6) | 9.4 ± 2.4 | 8.4 ± 1.7 | -1.0 (-3.2, 1.3) |
| | Control (N=21, n=5) | 6.6 ± 6.9 | 6.9 ± 1.9 | 0.3 (-1.3, 1.8) |

(A) Data are Mean ± SD for continuous variables. Outcome mean differences between pre- and during-intervention phases are shown with corresponding 95% confidence intervals.

Overall, sleep duration remained stable across all groups, with comparable pre- and during-intervention averages in both intervention groups relative to the control group. Physical activity outcomes showed modest changes: the intervention groups exhibited small declines in average daily flights climbed and step counts. For screen time, a slight reduction was observed in the goal-setting group, while the break and control groups showed minor increases.

## DISCUSSION

The MOVE@NUS pilot study demonstrated the feasibility of using a hybrid study design that combined continuous monitoring with multiple sequentially embedded randomized controlled trials among university students. The approach proved acceptable to participants, with sustained engagement among a subset of students, and successful data collection via both self-reports and passive tracking. Data completeness was strongest for passively captured measures of physical activity, while more participant-dependent measures, such as manual screen time screenshots and uploads, showed greater attrition. These findings support the potential of digital cohorts with embedded micro-trials as a scalable model for evaluating behavior change strategies in real-world settings.

Enrollment rate was modest, consistent with our previous experience recruiting from similar student cohorts and other digital health interventions targeting young adults with strict technological requirements [14,63–65]. In designing this pilot, we deliberately focused on the Apple operating system because iPhone ownership is high among university students in Singapore, ensuring smoother integration with Apple HealthKit and a higher likelihood of compatibility [65,66]. However, requiring both an iPhone and Apple Watch (Series 6/SE or newer) constrained recruitment and may limit generalizability to broader populations. These barriers highlight a common issue in digital health research [65,66], and future studies should consider both Android and Apple users or provide standardized study devices, although such approaches would entail greater resource demands for app development and potentially increase participant burden associated with carrying additional devices [8,68,69].

Our findings also revealed important insights into the hierarchy of data collection burden and participant compliance. Passive data collection approaches achieve higher participant compliance than methods requiring active engagement, even when the required active engagement is minimal [7,68]. Among passive measures, completeness was lower for sleep data compared with physical activity, largely due to participants’ non-wear of watches at night, making physical activity outcomes the most complete. However, the superiority of passive data collection was also undermined by technical challenges [70]. HealthKit’s dependence on periodic app activity and participants’ uninstalling the app after completing intervention phases reduced retention [70–72]. This highlights a critical consideration of more robust technical infrastructure for app developers, including seamless background synchronization, persistent data collection across reinstalls, and simplified monitoring interfaces [68,69,72]. Consistent with recent systematic reviews, our observations have highlighted that user engagement varies significantly based on the level of active participation required [32,68]. While general questionnaires remain valuable for structured baseline and follow-up assessments, they cannot capture behavioral changes in real time. Active approaches such as EMAs may provide complementary contextual insights but may not always achieve sufficient completion rates for primary outcome measurement in the long term [32,68,70,72,73]. Thus, future digital health interventions should prioritize technically robust passive monitoring as the backbone of continuous data collection, supported by questionnaires for comprehensive assessments and ecological momentary prompts that provide rich context without overburdening participants [7,32,65].

Participants generally reported high usability and comfort with the app setup and that they approached participation with motivation and confidence. These are factors known to maintain engagement, supporting the feasibility of smartphone-based health interventions in university students [35,71,74]. Feedback also revealed differences in acceptability across behavioral domains: sleep and physical activity nudges were perceived as more useful, relevant, and satisfactory than screen time interventions. Unlike the sleep and physical activity phases, which delivered fewer and more personalized notifications, the screen time phase relied on more frequent and repeated prompts to interrupt usage and was positioned last in the sequence of RCTs [32,68]. Open-text feedback also revealed that while reminders raised awareness and supported habit formation, over-frequent notifications could reduce satisfaction or inadvertently increase device use. Participants expressed preferences for more automated tracking, progress feedback, and incentive systems [42,49,75]. These findings align with behavior change frameworks, highlighting the importance of goal reinforcement, social features, and reward mechanisms for sustaining engagement [76–78]. Furthermore, participants enrolled for three consecutive phases showed greater attrition than those enrolled for two consecutive phases, suggesting an optimal engagement window of two to three months for intensive digital health interventions among students. This finding aligns with prior reports of high attrition in digital interventions, with studies reporting dropout rates approaching 50% [63,73], and suggests that study duration may be a critical factor in retention [72]. These patterns suggest that digital nudges may be more effective for specific behaviors for specific amounts of time, and underscore the need for adaptive, context-sensitive personalization to heterogeneous populations to account for baseline behaviors and individual preferences [49,75,76,79].

Moreover, the aim of this pilot was not to identify the single most effective intervention, but rather to validate the feasibility of a unified framework for delivering, personalizing, and optimizing multiple digital interventions within one platform. Although preliminary findings did not indicate substantial improvements in movement behaviors, the study demonstrated that the framework is feasible and practical for testing diverse formats and contents of digital nudges in naturalistic settings [10,32,79]. Considering the insufficient sleep and excessive screen usage [14,19,80], the trend toward increased sleep duration and reduced screen time suggests potential intervention effects that may become evident with larger sample sizes or longer intervention periods. The study also successfully trialed different approaches to personalizing EMIs, combining both passive tracking data and participant preferences, across multiple behavioral domains. In addition, we evaluated complementary methods for assessing intervention effects, including passive tracking and EMA responses, showing that these are feasible to be implemented within a single platform. Collectively, these findings provide proof of concept for the hybrid framework while emphasizing the need for more efficient study designs in future research. Although we used traditional RCT formats in this feasibility phase, alternative designs such as micro-randomized trials or factorial experiments may be more efficient for optimizing intervention components in subsequent larger-scale studies [81].

In summary, several design modifications should be considered to improve feasibility and scalability. Future trials may benefit from platform diversification (including both Apple and Android) to enhance recruitment, as well as strengthening technical infrastructure to ensure reliable passive data capture. For EMIs, greater emphasis should be placed on context-sensitive personalization that adapts to real-time behaviors and user preferences. Graduated engagement models such as milestone-based incentives and progress tracking, and an optimized intervention duration of two to three months, or several sequential phases of similar length with follow-up to refresh engagement and gather feedback, could help reduce participant burden and maintain retention. Finally, domain-specific tailoring is likely needed, as behaviors such as sleep, physical activity, and screen time may each necessitate distinct intervention approaches.

### Study limitations

Several limitations should be acknowledged. First, restricting eligibility to Apple users who own both an Apple smartphone and smartwatch limits the generalizability of findings to the broader university population and may introduce socioeconomic biases. Second, the relatively small sample size reduced statistical power to detect meaningful differences between groups. Third, due to discomfort wearing the Apple Watch overnight, many participants did not wear the device during sleep, leading to substantial data loss. As a result, time in bed was used as a proxy for sleep duration, which may overestimate actual sleep and limit the precision of sleep-related findings [74,82–84]. This emphasizes the need for future studies to carefully balance measurement comprehensiveness with participant comfort and burden when deploying wearable-based protocols [77,82,85]. Finally, since the intervention groups received daily nudges while the control groups received no intervention, participants in the intervention groups might perceive a greater level of observation. This awareness might unintentionally inflate positive responses in subjective, self-reported outcomes, which might introduce a risk of performance bias and the Hawthorne effect. While objective metrics from Apple HealthKit are less susceptible to this bias, the subjective findings were interpreted with caution, and the study prioritized feasibility assessment over definite effectiveness evaluation regarding this limitation.

## CONCLUSION

The MOVE@NUS pilot study demonstrates the feasibility of using a hybrid study design that combines continuous digital monitoring with three sequentially embedded RCTs among university students while also revealing important implementation challenges and design considerations. The purpose of this pilot was not to determine the most effective single intervention, but to validate the framework for delivering, personalizing, and optimizing multiple interventions within a single platform. Although strict inclusion criteria limited recruitment, the study demonstrated the potential for using personal smartphones and wearable devices to support healthy behavior change in naturalistic settings. Findings on acceptability differed across behavioral domains, optimal engagement duration, and technical implementation challenges, offering practical guidance for refining digital health interventions. Future studies can build on this foundation by refining intervention designs, diversifying platforms or providing devices to broaden participation, and considering alternative study designs such as micro-randomized or factorial trials to optimize efficiency. The insights regarding data collection hierarchy, retention patterns, and intervention acceptability will also inform the development of scalable, effective, and sustainable digital health interventions for emerging adults during this critical developmental period.

## DECLARATIONS

## Acknowledgements

We would like to thank the National University of Singapore University Health Centre for their significant contribution to participant recruitment, and all participants for their time and involvement in the study. We used the generative AI tool ChatGPT by OpenAI to assist in refining language expression, formatting, and overall readability after the manuscript was drafted. No AI tools were used for data collection, interpretation, or citation generation. All AI-assisted edits were carefully reviewed and verified by the authors, and all authors approved the final manuscript content.

## Data availability

Access to the final trial dataset is restricted to the principal investigator and designated members of the research team. Data has been stored securely, and any shared data will be anonymised and de-identified to ensure participant confidentiality. Vendor access (e.g., Apple, REDCap) is strictly governed by institutional Data Use Agreements. For additional information, please contact the lead author.

## Author contributions

MC, NP, SE, and FMR conceptualized and designed the MOVE@NUS study. MC, XHC, and MM were responsible for the operational conduct and implementation of the study. XHC, MM, ST, SZ, KJ, TT, NP, SE, and FMR participated in the in-house testing of the developed app and provided feedback for refinement. SE and FMR supervised the study and provided critical revisions to enhance the scientific rigor of the protocol manuscript. MC drafted the initial manuscript, with all authors reviewing and approving the final version for submission.

## ABBREVIATIONS

CI: Confidence Interval
EMA: Ecological Momentary Assessment
EMI: Ecological Momentary Intervention
VPA: Vigorous Physical Activity
PA: Physical Activity PRs: Permanent Residents
REDCap: Research Electronic Data Capture
SD: Standard Deviation
SGD: Singapore Dollar

